# A preliminary study of commercially available general-purpose chest radiography artificial intelligence-based software for detecting airspace opacity lesions in COVID-19 patients

**DOI:** 10.1101/2021.12.22.21268176

**Authors:** Munemura Suzuki, Aruta Niimura, Yusuke Nakamura, Yujiro Otsuka

## Abstract

**Purpose:** To validate commercially available general-purpose artificial intelligence (AI)-based software for detecting airspace opacity in chest radiographs (CXRs) of COVID-19 patients.

**Materials and Methods:** We used the ieee8023-covid-chestxray-dataset to validate commercial AI software capable of detecting “Nodule/Mass” and “Airspace opacity” as regions of interest with probability scores. From this dataset, we excluded computed tomography images and CXR images taken using an anteroposterior spine view and analyzed CXR images tagged with “Pneumonia/Viral/COVID-19” and “no findings.” A radiologist then reviewed the images and rated them on a 3-point opacity score for the presence of airspace opacity. The maximum probability score of airspace opacity for each image was calculated using this software. The difference in each maximum probability for each opacity score was evaluated using Wilcoxon’s rank sum test. The threshold of the probability score was determined by receiver operator characteristic curve analysis for the presence or absence of COVID-19, and the true positive rate (TPR) and false positive rate (FPR) were determined for the individual and overall opacity scores.

**Results:** Images from 342 patients with COVID-19 and 15 normal images were included. Opacity scores of 1, 2, and 3 were observed in 44, 70, and 243 images, respectively, of which 33 (75%), 66 (94.2%), and 243 (100%), respectively, were from COVID-19 patients. The overall TPR and FPR were 0.82 and 0.13, respectively, at an area under the curve of 0.88 and a threshold of 0.06, while the FPR for opacity score 1 was 0.18 and the TPR for score 3 was 0.97.

**Conclusion:** Using a public database containing CXR images of COVID-19 patients, commercial AI software was shown to be able to detect airspace opacity in severe pneumonia.

**Summary:** Commercially available AI software was capable of detecting airspace opacity in CXR images of COVID-19 patients in a public database.

## Introduction

Coronavirus disease 2019 (COVID-19) has spread worldwide. This pandemic caused a crisis-level shortage of hospital beds in many countries. In Japan, the number of positive cases per day reached 25,851 (20.6/100,000) on August 25, 2021, and the hospitalization rate decreased to 11.8%^1^. Therefore, there is an urgent need to establish a method for the appropriate allocation of medical resources.

Chest radiography (CXR) is expected to be an efficient triage tool for COVID-19 due to its low operational cost and high throughput. Although it is less sensitive than computed tomography (CT) for early detection of pneumonia^2^, it has been reported to be sufficiently sensitive for detecting severe disease and high-risk CXR findings such as bilateral opacities, multifocal opacities, and upper or middle zone opacities^3^. Moreover, artificial intelligence (AI)-based patient prognostic models have been reported^4,5^ and are expected to prove useful for clinical triage and workflow optimization.

In this study, we hypothesized that commercially available general-purpose AI software for CXR could detect airspace opacity in COVID-19 patients. As an initial study prior to a clinical study, we used the publicly available COVID-19 CXR database^6^ to validate the software and examine its performance.

## Materials and Methods

We used an anonymized public dataset^6^ that has already been widely used for AI research^7,8^ on COVID-19. In accordance with the ethical guidelines for medical and health sciences research involving human subjects^10,11^, no review by an institutional review board was required.

Because the dataset contains both CT and CXR images, we first extracted only CXR images. Second, we filtered suspected or confirmed COVID-19 positive patients and patients with no findings as controls. Third, we excluded images that tagged “AP supine views” because we were more interested in detecting imaging findings in asymptomatic to moderately ill patients than in detecting findings in images of patients with pre-existing severe pneumonia. Moreover, we excluded lateral-view images.

A radiologist with 17 years of experience independently rated the presence of airspace opacity on a 3-point scale based on confidence levels. The criteria were as follows: opacity confidence score 1, negative; opacity confidence score 2, indeterminate (including ground-glass opacity and suboptimal image quality); opacity confidence score 3, definite airspace opacity. In this preliminary study, we did not consider the zones containing shadows, the number of shadows, or the size of the shadows.

We used commercially available AI software (Plus.CXR; Plusman LLC, Tokyo, Japan; Pharmaceuticals and Medical Devices Agency certification cleared) that requires anteroposterior (AP) or posteroanterior (PA) CXR images as input and outputs the regions of interest (ROIs) that indicate “Nodule/Mass” class or “Airspace Opacity” class and their probability scores. In addition to the coordinates of the ROIs and the probability of the class, we could identify the CXR zones [right and left side of upper (from the apex to the inferior border of the second rib), middle (from the second to fourth rib), and lower (from the fourth rib to the inferior border of the lung) zones] where the ROIs exist. We also measured the cardiothoracic ratio as a reference value. Figure 1 shows a CXR image overlaid with the AI output results. Note that the threshold settings used for the display were disabled. The bounding box shows the name of the detected class, the probability score, and the zone where the shadow exists. Figure 1a shows an image with an opacity confidence score of 1 for visual evaluation, showing a pale infiltration to the frosted shadow. Figure 1b shows an image with an opacity confidence score of 2, which shows a dense airspace opacity shadow with bilateral upper lung zone predominance.

**Figure 1.**
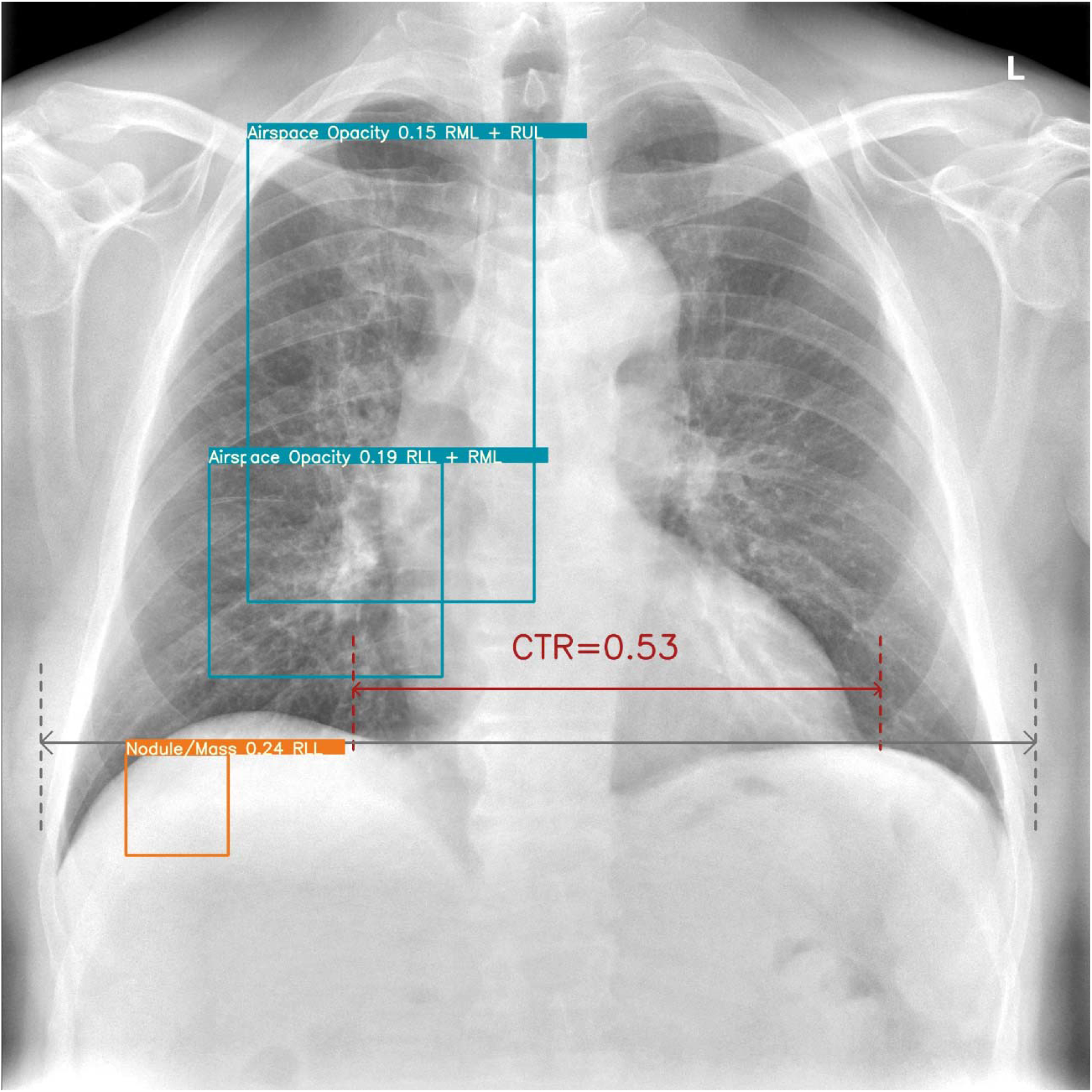
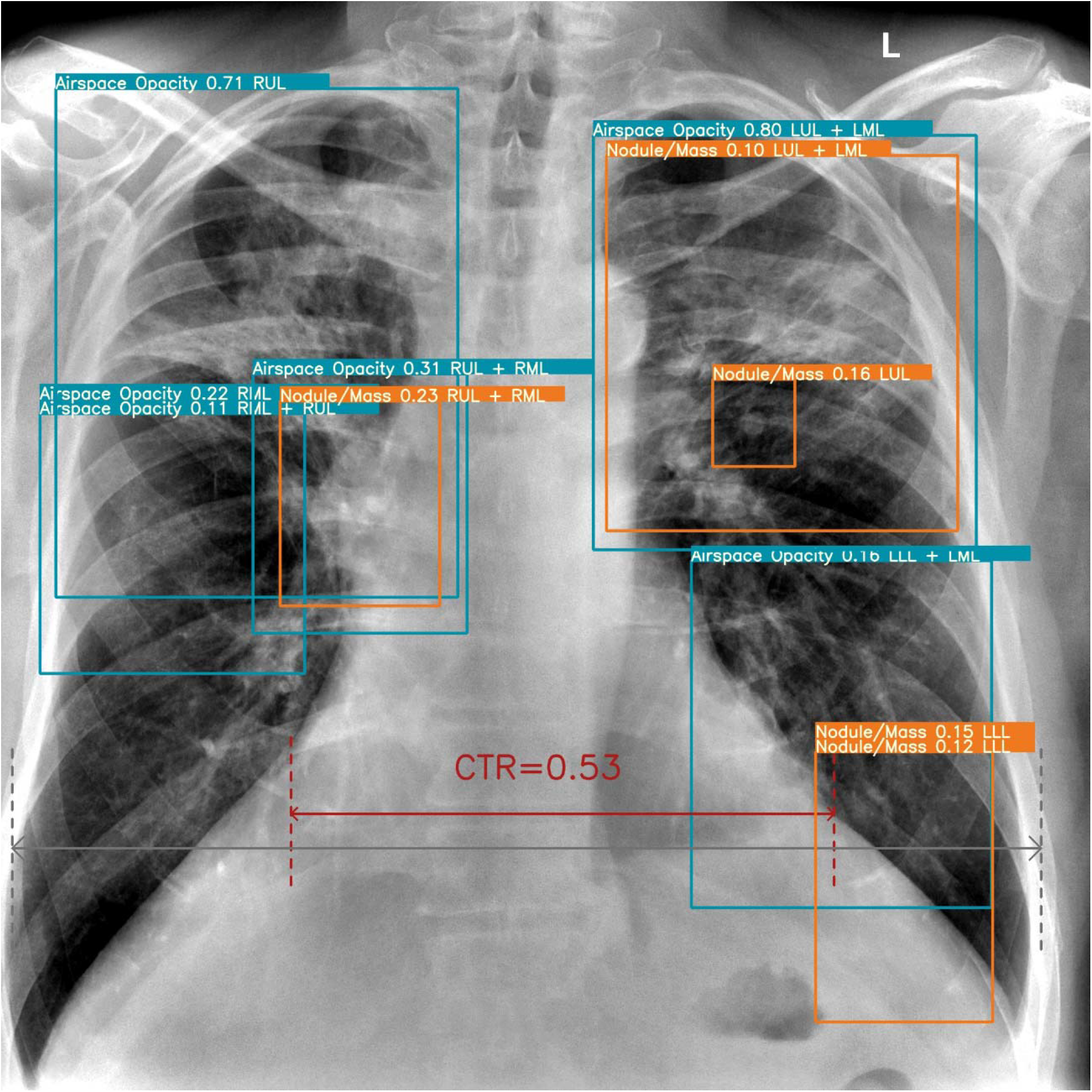
Examples of chest radiography (CXR) images of patients with COVID-19 overlaid with the artificial intelligence output results. Note that the threshold setting used for the display has been disabled. The bounding box shows the name of the detected class, the probability score, and the zone (right and left sides of the upper, middle, and lower zones) where the shadow exists. The cardiothoracic ratio (CTR) as a reference value is also shown. Figure 1a shows pale airspace opacity to ground glass opacity in the right upper to lower zone. Figure 1b shows dense airspace opacity with collapse and pale airspace opacity to ground glass opacity in both lungs. Some “Nodule/Mass” classes were also detected with a relatively low probability. doi: 10.6084/m9.figshare.1227500966, CC BY 3.0.

Statistical analyses were performed using Python 3.6, Numpy (version 1.18.1), Pandas (version 1.0.2), scikit-learn (version 0.22.2. post 1), and Scipy (1.4.1). The probability scores of all images and images with each opacity score were reported as medians and interquartile ranges (IQRs).

We assessed the differences in probability scores using Wilcoxon’s test. P values of ≤0.05, were considered statistically significant. Receiver operator characteristic (ROC) curves were generated from the presence or absence of COVID-19 and probability scores for all images, and the area under the ROC curve (AUC) and threshold values were determined. The true positive rate (TPR) and the false positive rate (FPR) for the individual and overall capacity scores were calculated using the calculated thresholds.

## Results

Of the 950 images in the dataset, 866 were CXRs; from 504 COVID-19 and 18 normal images, 342 COVID-19 images and 15 normal images were finally available for analysis. Figure 2 shows the image selection flowchart and its breakdown.

**Figure 2.**
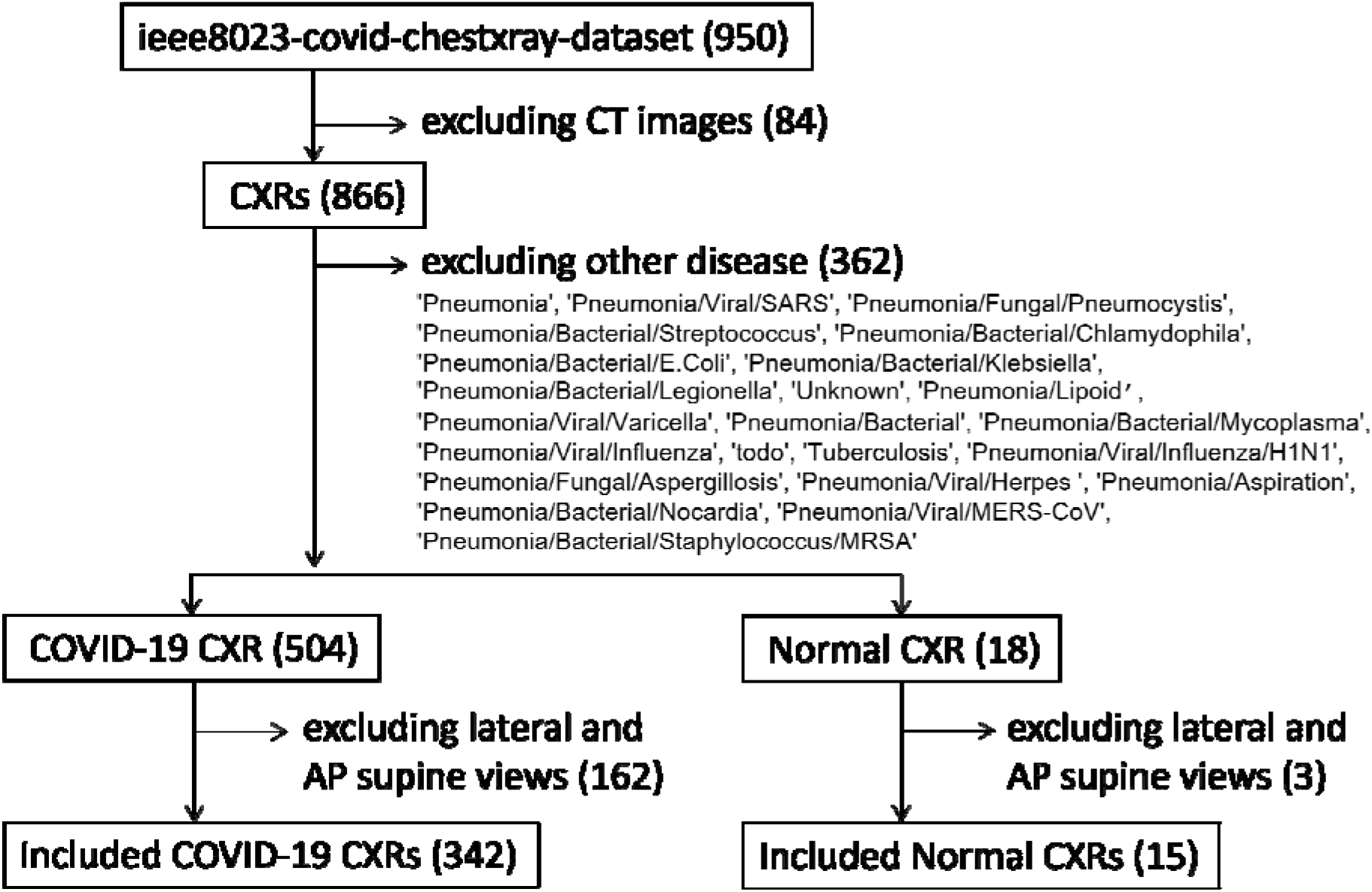
Flowchart for selecting images from a database.

Opacity confidence scores of 1, 2, and 3 were found in 44, 70, and 243 images, respectively, of which (75%), 66 (94.2%), and 243 (100%), respectively, were from patients with COVID-19. Figure 3 shows a box-and-whisker plot of the results of the quantitative and probability scores. The medians for overall and opacity confidence scores of 1, 2, and 3 were 0.35 (IQR: 0.11-0.55), 0.02 (IQR: 0.00-0.03), 0.11 (IQR: 0.04-0.25), and 0.47 (IQR: 0.33-0.59), respectively. There was a significant difference between the probability scores of confidence scores 1, 2, and 3 (p < 0.001).

**Figure 3.**
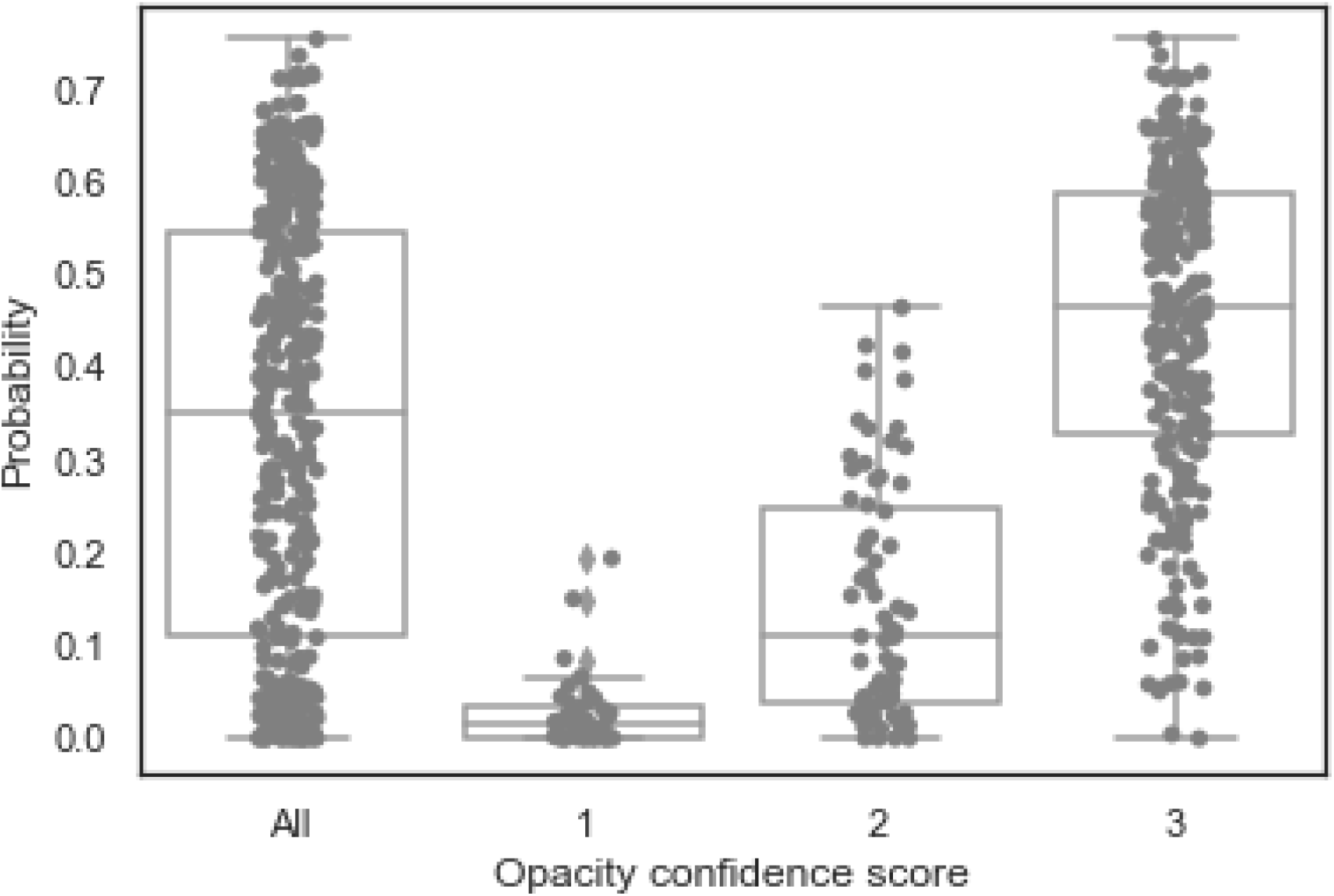
The maximum value of probability for each image obtained by the artificial intelligence model was differed significantly among the opacity confidence scores of 1, 2, and 3 based on visual assessment (p < 0.001). The box in the boxplot shows the median and interquartile range (IQR), with the whiskers extending to points within 1.5 IQR of the IQR boundary. Gray dots indicate data points, and diamonds indicate outliers.

Figure 4 shows the ROC curve generated for the diagnosis of COVID-19 using all images. The AUC was 0.88. Figure 5 shows the heat map of the confusion matrix for all images and for those with scores of 1, 2, and 3 when the threshold was set to 0.06. The TPR and FPR for each score are shown in Table 1.

**Figure 4.**
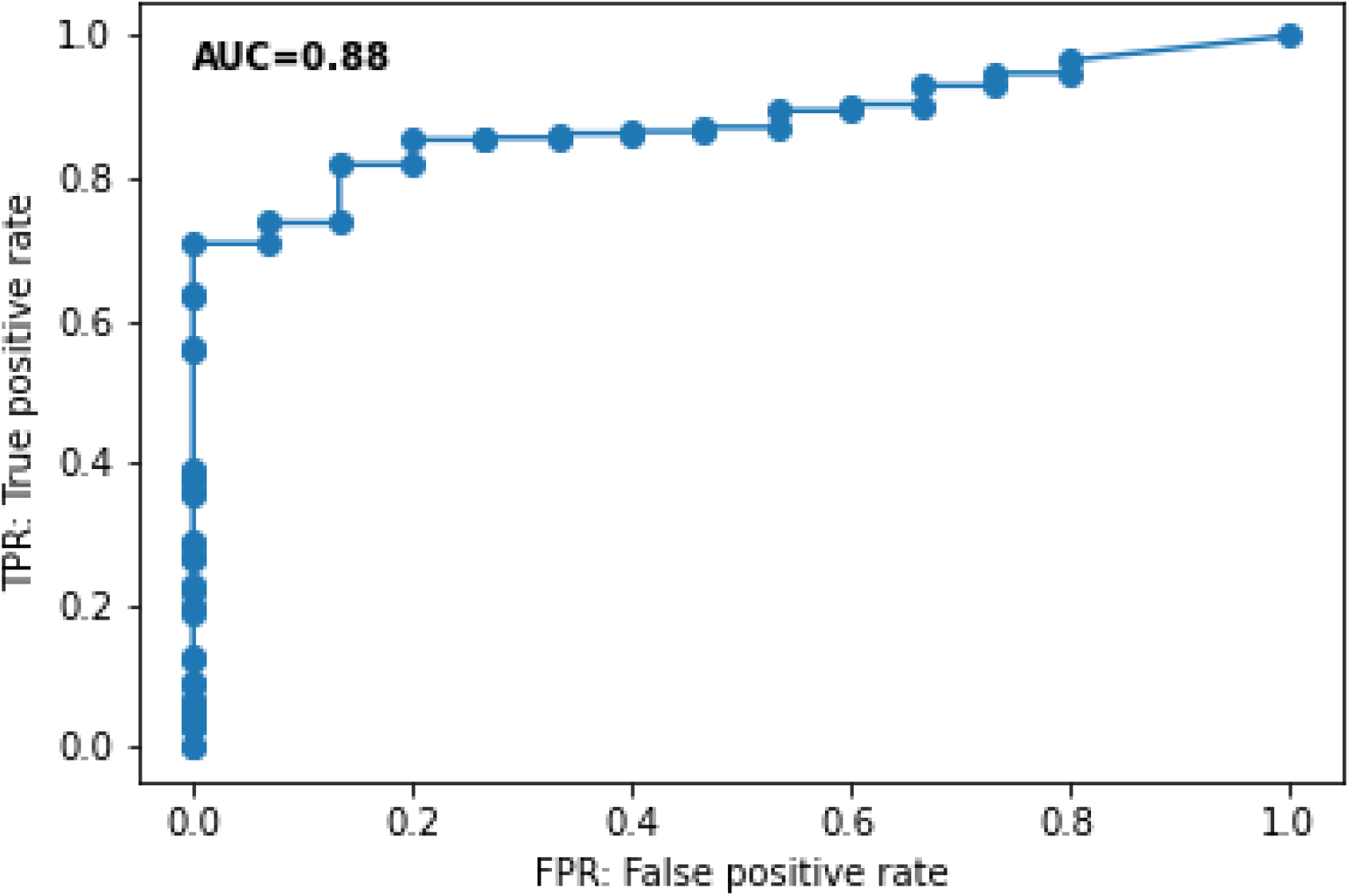
Receiver operating characteristic curve shows the performance of the probability obtained with artificial intelligence for predicting COVID-19. AUC: area under the curve.

**Figure 5.**
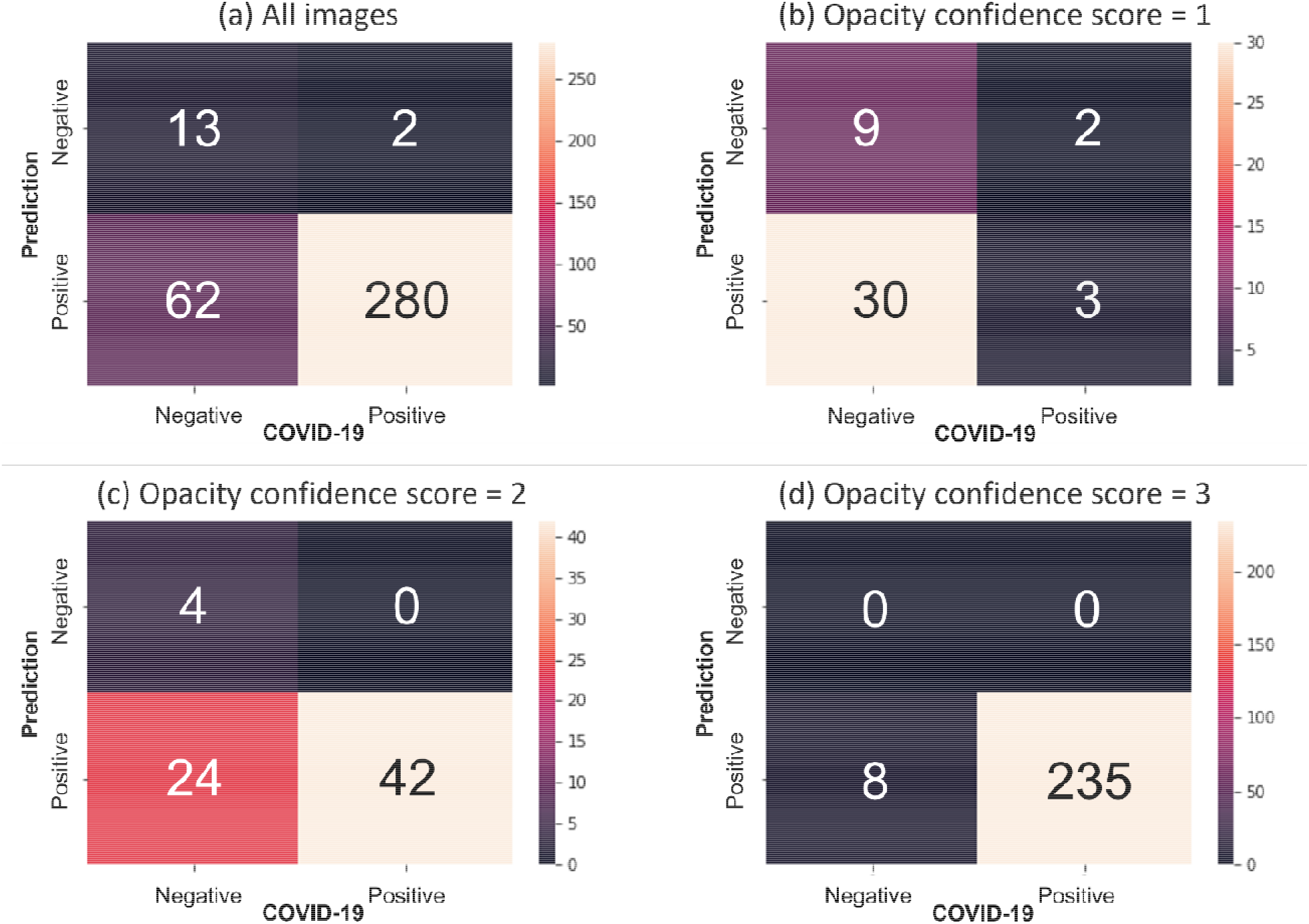
Heat maps of the confusion matrices for all images and each opacity confidence score are shown. The matrix in each figure is as follows: top left, true negative; bottom left, false positive; top right, false negative; bottom right, true positive. The numbers in the matrix indicate the number of images in each block.

**Table 1.** TPR and FPR for all images and the images of opacity confidence score 1, 2, 3, respectively (threshold of probability score = 0.06)

|  | All images | Opacity confidence score |  |  |
| --- | --- | --- | --- | --- |
|  | (n = 357) | 1 (n = 44) | 2 (n = 70) | 3 (n = 243) |
| TPR | 0.82 | 0.09 | 0.63 | 0.97 |
| FPR | 0.13 | 0.18 | 0.0 | 0.00 |

Abbreviations: FPR, false positive rate; TPR, true positive rate.

## Discussion

During the COVID-19 pandemic, many countries experienced a depletion of medical resources, whose efficient distribution has become a challenge. Although CXR is reported to have a lower initial diagnostic sensitivity for COVID-19 than that associated with CT^2^, it is widely used for patient monitoring and screening because of its advantages such as the equipment’s low cost and simplicity, transportability to the bedside, short imaging time, and ease of decontamination. In this study, we showed that COVID-19 lung lesions can be detected using commercially available AI software. We discuss whether this model can be used for efficient COVID-19 practice and the additional research needed to use it.

In this study, we showed that the presence of airspace opacity can be evaluated using the maximum probability score calculated by our model. The results also suggest that our model is consistent with the qualitative evaluation of shadows based on visual assessment by radiologists. Liang et al. analyzed prognostic factors, including the presence of CXR and CT abnormalities, as imaging information along with clinical information and the severity and reported that CXR was a statistically significant independent prognostic factor^8^. Other reports have revealed that the CXR severity score, calculated from the CXR shadow quality and shadow coverage, is a predictor of hospital admission^12^, intubation^8,12^ oxygen administration^3,13^, and death^8,14^. Sasaki et al. compared consolidation with ground-glass attenuation (GGA) scoring and reported that GGA scoring was not a predictor of oxygen administration1313. Considering the learning cost for raters to perform uniform image evaluation and the human cost and time cost for reading images, the use of AI-based evaluation criteria is considered worth exploring.

There are two possible strategies for creating an AI-based, image-based prognostic model. The first is to create a model that uses the severity scores provided by radiologists for learning. Li et al. developed a model to calculate the pulmonary radiography severity score (PXS) using the radiologist’s CXR-finding score as a reference standard and showed that the PXS correlated with the occurrence of intubation or death within three days44. The other is to create a model that learns prognostic features from the image itself. Jiao et al. reported a prognostic model differentiating critical (induction of ventilation, intensive care unit admission, or death) from non-critical disease, using imaging and clinical data as their inputs55. They also reported that the addition of clinical information improved the accuracy of prognostication. Our model adopted the former approach, and we believe that this approach is preferable during the COVID-19 pandemic.

We believe that there are two advantages to a model that uses radiologist-derived severity scores for learning. First, they can be “explainable AI.” Because the interpretation of AI output results is ultimately the responsibility of humans, the task of image review by clinicians remains. Second, it is easy to create new models using existing models in a situation where factors that affect the clinical course, such as vaccination, antibody cocktail therapy, and mutant strains with different toxicities, emerge consecutively. It is expected that a model for extracting one-stop prognostic features from CXR using AI will need to be developed for each cohort. Conversely, it is relatively easy to create a new prognostic model by varying the threshold value of the model that extracts the probability and area of findings from images.

This preliminary study had several limitations. First, we used public data. The image quality varied, and there were some missing clinical data. In fact, the team that created this database advises not to use them to determine diagnostic performance^15^. However, because we used public data not used for training for validation, the impact of database bias^4^ is expected to be small. Second, images were evaluated by a single person. Although there is interobserver variability in the diagnosis of pneumonia^16^, in this study, each group of confidence scores by a single radiologist could be separated by AI-based probability scores with significant differences. The clinical significance of the overlap is unclear, but it is a potential consideration for future research. Third, we did not evaluate all the lesions extracted by our model. This is because there is no gold standard such as CT, and it is difficult to perform a detailed evaluation because of differences in the image quality and size compared to those of DICOM format images for diagnosis. This is an issue for consideration in future research.

In conclusion, our hypothesis that commercially available AI-based software could detect opacity regions in CXRs of COVID-19 patients was proved. We hope to conduct clinical research and develop a prognostic model using the output results of this AI to help in the treatment of COVID-19 infections.

## Data Availability

All data produced in the present study are available upon reasonable request to the authors

